# Validation of the performance of a point of care molecular test for leprosy: from a simplified DNA extraction protocol to a portable qPCR

**DOI:** 10.1101/2024.02.29.24303527

**Authors:** Amanda Bertão-Santos, Larisse da Silva Dias, Marcelo Ribeiro-Alves, Roberta Olmo Pinheiro, Milton Ozório Moraes, Fernanda Saloum de Neves Manta, Alexandre Dias Tavares Costa

**Affiliations:** Laboratório de Ciências e Tecnologias Aplicadas à Saúde (LaCTAS), Instituto Carlos Chagas, Fundação Oswaldo Cruz (FIOCRUZ), Curitiba, Brazil; Laboratório de Hanseníase, Instituto Oswaldo Cruz, Fundação Oswaldo Cruz (FIOCRUZ), Rio de Janeiro, Brazil; Laboratório de Pesquisa Clínica em DST e AIDS, Instituto Nacional de Infectologia Evandro Chagas, Fundação Oswaldo Cruz (FIOCRUZ), Rio de Janeiro, Brazil

**Keywords:** diagnostic test, leprosy, point-of-care, DNA extraction, portable qPCR

## Abstract

The study aimed to optimize qPCR reactions using oligonucleotides from the first Brazilian molecular diagnostic kit for leprosy on a portable platform (Q3-Plus). In addition, we sought to develop a simplified protocol for DNA extraction that met point-of-care criteria. During optimization on the Q3-Plus, optical parameters, thresholds, and cutoffs for the *16S rRNA* and RLEP targets of *M. leprae* were established using synthetic DNA, purified DNA from *M. leprae*, and pre-characterized clinical samples. In the simplified extraction step, different lysis solutions were evaluated using chaotropic agents, and purification was carried out by depositing the lysed material on FTA cards. The complete protocol (simplified extraction + qPCR on the portable platform) was evaluated with pre-characterized clinical skin biopsy samples and compared with standard equipment (QuantStudio-5). LOD_95%_ for the optimized reactions was 113.31 genome-equivalents/μL for *16S rRNA* and 17.70 genome-equivalents/μL for RLEP. Among the lysis solutions, the best-performing was composed of urea (2 M), which provided good dissolution of the skin fragment and a lower Ct value, indicating higher concentrations of DNA. The complete technological solution showed a sensitivity of 52% in reactions. Our results highlight the need for additional optimization to deal with paucibacillary samples, but also demonstrate the applicability of the portable platform in the detection of *M.* leprae in low infrastructure settings.

## Introduction

Leprosy is a chronic and progressive infectious disease with worldwide distribution caused by *Mycobacterium leprae* or *M. lepromatosis* (HAN et al., 2008). It presents tropism for peripheral nerves and skin, although other organs might also be affected. Progression of the disease might cause deformities and different degrees of physical disability (FI). Given its signs and symptoms, leprosy can be manifested in a broad spectrum of clinical forms, some of which often lead to misdiagnosis with other dermatological, osteoarticular, or neurological conditions, and even other diseases (NEVES et al., 2023; DHARMAWAN et al., 2022).

Early diagnosis is essential for proper treatment and control of the disease’s clinical progression and community transmission (STEINMANN et al., 2017). Due to its inability to grow *in vitro*, direct diagnostic techniques such as culture and isolation are not feasible (MACIEIRA, 2000). Diagnosis is usually late or non-existent because it is mainly based on the patient’s clinical and epidemiological information, guided by anamnesis and physical examination, which demands physician’s expertise (WHO, 2022; HENRY et al., 2016). In the absence of a gold standard diagnostic test, complementary tests are often employed, such as histopathology and bacilloscopy of the slit-skin smear. These tests, however, exhibit variations in sensitivity accordingly the clinical form of the disease and rely on experience for contextualization and interpretation (NEVES et al., 2023; WHO, 2022; HENRY et al., 2016).

Molecular tests, particularly real-time polymerase chain reaction (qPCR), are sensitive and specific, contributing to the early identification of various pathologies (MADAMET et al., 2022; RAMPAZZO et al., 2022; YU et al., 2021). Although molecular detection tests are available, the diagnosis of leprosy continues to rely primarily on clinical diagnosis (WHO, 2022). This is due to the variations in clinical presentations, especially in multibacillary and paucibacillary cases, as the analytical sensitivity of tests varies according to the bacillary load of the infection (BARBIERI et al., 2019).

Recently, Brazilian health authorities (ANVISA) granted registration for the first national qPCR NAT Hans kit (IBMP, Brazil), developed by the Oswaldo Cruz Foundation (Fiocruz/RJ) (MANTA et al., 2022). This kit specifically targets the genetic markers *16S rRNA* and RLEP *M. leprae*, demonstrating sensitivity and specificity of 91% and 100%, respectively. Additionally, it utilizes human *18S rRNA* as an internal control, ensuring proper DNA extraction. The incorporation of this molecular test into the routine of healthcare professionals engaged in active case finding for leprosy can overcome the intrinsic limitations of direct diagnostic methods such as bacilloscopy, histopathology, and indirect serology tests, thereby expanding the detection capacity, especially for the paucibacillary (PB) clinical form (BARBIERI et al., 2019).

Tools that assist in population screening play a crucial role in active case finding and early diagnosis of leprosy. Screening tests exhibit higher sensitivity, meaning they can more accurately identify positive cases for the disease of interest. A positive result in a screening test should guide the patient toward further assessments that allow for an accurate diagnostic investigation, as is done in the case of leprosy (WHO, 2020). However, the implementation of leprosy molecular detection tests in the field faces limitations. The requirement for thermolabile reagents and robust equipment hinders access to diagnosis in remote and low-infrastructure regions. Furthermore, the method requires prior DNA extraction, demanding investments in costly commercial kits and time-consuming to obtain the sample (MANTA et al., 2020; ALI et al., 2017; WANG et al., 2016). Regarding nucleic acid testing (NAT) based diagnosis, the primary challenges are related to the pre-analytical phase, including specimen collection and biological material extraction. The need for sensitive instruments, such as centrifuges, and high-cost reagents, as well as the proper disposal of the chemical residues generated in these steps, represents the primary limitations for their applicability in resource-limited settings (ALI et al., 2017; DINEVA et al., 2007).

The aim of the study was optimizing qPCR reactions using the NAT Hans kit (IBMP, Brazil) on the portable Q3-Plus instrument, while concurrently developing a simplified DNA extraction protocol for skin samples, aiming the detection of *M. leprae* DNA. The resulting prototype enables the implementation of a leprosy screening test in remote areas, facilitating the active search of positive cases and monitoring by health authorities responsible for underserved populations.

## Materials and Methods

### Ethics Statement

The present research project was approved by the Ethics Committee of the Oswaldo Cruz Institute (CAAE: 52565521.2.0000.5248, number: 5.131.588 approved on November 26, 2021). Suspected leprosy patients attending the Souza Araújo Out-Patient Unit (ASA), a leprosy reference center from Brazilian Ministry of Health at Oswaldo Cruz Institute – Fiocruz – RJ – Brazil, provided written consent to participate in the project. In the case of minors, formal written consent was obtained from the patient’s guardian. The selection of samples was carried out according to the occurrence of cases attended at the clinic.

### Synthetic DNA

Synthetic double-stranded DNA (gBlock, IDT, USA) containing the sequences of genomic markers for *M. leprae* (*16S rRNA* and RLEP) and the human genomic marker (*18S rRNA*) was used for the optimization of qPCR reactions on the portable equipment (Q3-Plus). Paired evaluations were also performed on the standard equipment (Quantstudio 5 – QS-5). The lyophilized synthetic DNA was constituted at 10 fg/µL (equivalent to 10^4^ copies/µL). To obtain the sample at 10^5^ copies/µL, the gBlock was amplified by qPCR, and its product was purified using the QIAquick PCR Purification Kit (Qiagen, Germany). The concentration was determined by interpolating Cycle threshold (Ct) values on the standard curve. When necessary, samples were diluted in Tris-EDTA (TE) buffer (pH 8.0) for standard curve analyses.

### M. leprae cells

*M. leprae* cells (Thai-53, at 10^6^ cells/mL) obtained from nude mice footpads were kindly provided by Dr. Patricia Sammarco Rosa from Lauro de Souza Lima Institute (Bauru, São Paulo, Brazil).. The cells were diluted at 1:10 in Tris-EDTA (TE) buffer (pH 8.0) for the construction of the standard curve.

### Determination of optical parameters, threshold, and cutoff in the optimization of the portable equipment: Q3-Plus

For the determination of optical parameters corresponding to readings through the FAM, VIC, and ROX channels on the Q3-Plus equipment, different values of exposure time, gain, and light intensity were evaluated (TABLE 1). The alterations aimed to improve the sensitivity of the reactions and achieve a good fluorescence amplitude. To achieve this, various concentrations (10^4^ – 10^0^ copies/µL) of synthetic DNA were evaluated using qPCR on the portable platform, and based on reaction efficiency and amplification curve characteristics, the parameters were assessed.

The baseline was automatically defined by the instrument’s software (Q3-Plus V2 Suite, version 4.0, ST Microelectronics). The threshold for each target was manually set by the operator through observation of fluorescence amplitude patterns and images captured by the software during reaction cycles with different concentrations of synthetic DNA, purified *M. leprae* DNA, pre-characterized clinical samples, as well as negative controls.

**TABLE 1.** – Optical parameters, including exposure time, gain and LED power, were assessed for reading analyses using the Q3-Plus equipment. *FAM – Probe to target 16S rRNA of M. leprae; VIC – probe to target RLEP of M. leprae; ROX – Probe to target 18S rRNA of mammals.

| FAM |  |  | VIC |  |  | ROX |  |  |
| --- | --- | --- | --- | --- | --- | --- | --- | --- |
| Exposure Time (s) | Gain | LED Power | Exposure Time (s) | Gain | LED Power | Exposure Time (s) | Gain | LED Power |
| 1 | 14 | 5 | 1/2 | 11 | 5 | **1** | **15** | **7** |
| 1 | 14 | 7 | 1/2 | 15 | 8 | - | - | - |
| **1** | **14** | **8** | 1/2 | 15 | 9 | - | - | - |
| 1 | 15 | 7 | 1/2 | 15 | 10 | - | - | - |
| 1 | 15 | 10 | 1 | 14 | 5 | - | - | - |
| - | - | - | **1** | **14** | **9** | - | - | - |
| - | - | - | 1 | 15 | 8 | - | - | - |
| - | - | - | 2 | 13 | 10 | - | - | - |
| - | - | - | 2 | 14 | 7 | - | - | - |
| - | - | - | 2 | 14 | 9 | - | - | - |
| - | - | - | 2 | 15 | 7 | - | - | - |
| - | - | - | 2 | 15 | 10 | - | - | - |
| - | - | - | 2 | 16 | 10 | - | - | - |

The establishment of the Ct value for the cutoff was conducted in the final stage of the study. For this purpose, two groups of pre-characterized clinical samples were analyzed: (i) The “standard” group consisted of skin biopsy samples extracted using a standardized commercial kit; (ii) The second group consisted of samples extracted using the protocol developed in the present study.

A Bland-Altman analysis was performed (ALTMAN & BLAND, 1983). The mean variations on Ct values to different equipment for the targets were added to the values of cutoff already established in the NAT Hans kit (IBMP, Brazil).

### Standard curve and analytical sensitivity on Q3-Plus and Quantstudio-5 equipment

The efficiency and determination of the detection limit for the qPCR reactions on the portable Q3-Plus instrument were obtained through a standard curve with logarithmic scale dilutions using synthetic DNA or purified *M. leprae* DNA (referred to as equivalent-genomes). The efficiency calculation for the reactions was done by substituting the slope value of the linear regression line into the efficiency formula (E=(10(–1/slope)-1).*100), following the MIQE Guidelines (BUSTIN *et al*., 2009).

The number of equivalent genome copies was estimated by interpolating Ct values obtained from the analysis of purified *M. leprae* DNA using the equation established after linear regression of the curve performed with synthetic DNA, considering known concentrations and the number of copies (10 fg/µL equivalent to 10^4^ copies/µL of synthetic DNA). For the analysis of the analytical sensitivity of the reaction, seven dilution points were considered, with a higher number of replicates (nine or ten) for the lower concentrations (BURD, 2010).

As a reference, identical analyses were performed using the standard Quantstudio-5 (QS-5) equipment.

### Reproducibility and Repeatability of qPCR reactions using synthetic DNA on the portable equipment

The reproducibility of the reaction was carried out by the independent operators over three consecutive days for concentrations ranging from 10^4^ to 10^0^ copies/µL. Subsequently, for three consecutive days, the same operators performed three reactions with lower concentrations ranging from 10^1^ to 10^0^ copies/µL. This division was due to the inherent limitation of the number of reactions that can be analyzed per chip (six wells). For each replicate, a new dilution was performed using synthetic DNA as the sample and Tris-EDTA buffer (pH 8.0) as the diluent (BURD, 2010).

### Clinical Samples

The clinical samples obtained in the present study were collected based on the occurrence of attendance at the Souza Araújo Clinic from the Oswaldo Cruz Foundation (Rio de Janeiro, Brazil). Skin biopsy collections were performed using a 3 mm surgical punch and stored in 70% ethanol until sample processing. Following clinical assessment, the samples were characterized according to established protocols, including clinical evaluation, histopathology, bacilloscopy, and singleplex qPCR (*16S rRNA*). Cases were classified as multibacillary (MB), paucibacillary (PB), or other dermatoses (OD) following WHO guidelines (WHO, 2018).

This study utilized 115 clinical skin biopsy samples, comprising 41 MB, 25 PB, and 49 OD (Supplemental Table S1). Of these, DNA was extracted from 62 skin biopsy samples (27 MB, 16 PB, and 19 OD) using commercial DNAeasy Blood and Tissue (Qiagen, Germany) and, 53 skin biopsy samples from leprosy patients and suspects (14 MB, 9 PB, and 30 OD) were extracted using both commercial DNAeasy Blood and Tissue (Qiagen, Germany) and the simplified DNA extraction protocol.

For the optimization of qPCR reactions on the portable equipment (Q3-Plus), exclusively samples extracted by the commercial protocol (Qiagen, Germany) were used.

### Standard methods used as guides for the methods developed

As a guide method for the protocols developed in the present study, clinical samples were also subjected to DNA extraction using a commercial kit. qPCR analyses using the NAT Hans kit were performed on a standard qPCR instrument (QuantStudio-5).

### Commercial DNA Extraction

DNA extraction from skin biopsy samples (3 mm) stored in 70% alcohol was performed using DNAeasy Blood and Tissue^®^ extraction kit (Qiagen, Germany) according to the manufacturer protocol. DNA concentration was estimated using NanoDrop^®^ (Thermo-Fisher Scientific, Waltham, MA, USA) and immediately stored at –20 °C.

### Experimental conditions for multiplex qPCR (NAT Hans) in standard equipment: QuantStudio 5

Detection of the two *M. leprae* targets (*16S rRNA* and RLEP) and the human internal control (*18S rRNA*) was performed as previously described by MANTA et al. (2022). Reactions were performed in a standard benchtop instrument (QuantStudio-5, Applied Biosystems, USA). Reactions were performed using the NAT Hans kit and were analyzed as manufacturer’s instructions. Reactions’ final volume was 25 μL, containing 5 μL of DNA. Cycling conditions used in QS-5 were 95 °C for 10 minutes, 45 cycles of 95 °C for 15 seconds, and 60 °C for 1 minute.

## Development of DNA extraction protocol and evaluation of the qPCR reactions in Q3-Plus

### Evaluation of lysis solutions utilizing the porcine skin model

Due to limitations regarding the availability of clinical samples from leprosy and suspected leprosy patients, porcine skin was used as a comparative model for the evaluation of six different solutions of lysis to DNA extraction protocols (TABLE 2) (HWANG et al., 2021; SUMMERFIELD et al., 2015). The evaluations were established in two ways: (i) visual observation regarding the dissolution/reduction of the skin fragment and changes in solution turbidity, and (ii) through qPCR for amplification of the mammalian *18S rRNA* gene (MANTA et al., 2020), as an analysis of the efficiency of each protocol. For the extraction of negative control, a pig skin fragment incubated with nuclease-free water was used, which underwent the same processing protocol as the other samples.

**TABLE 2.**
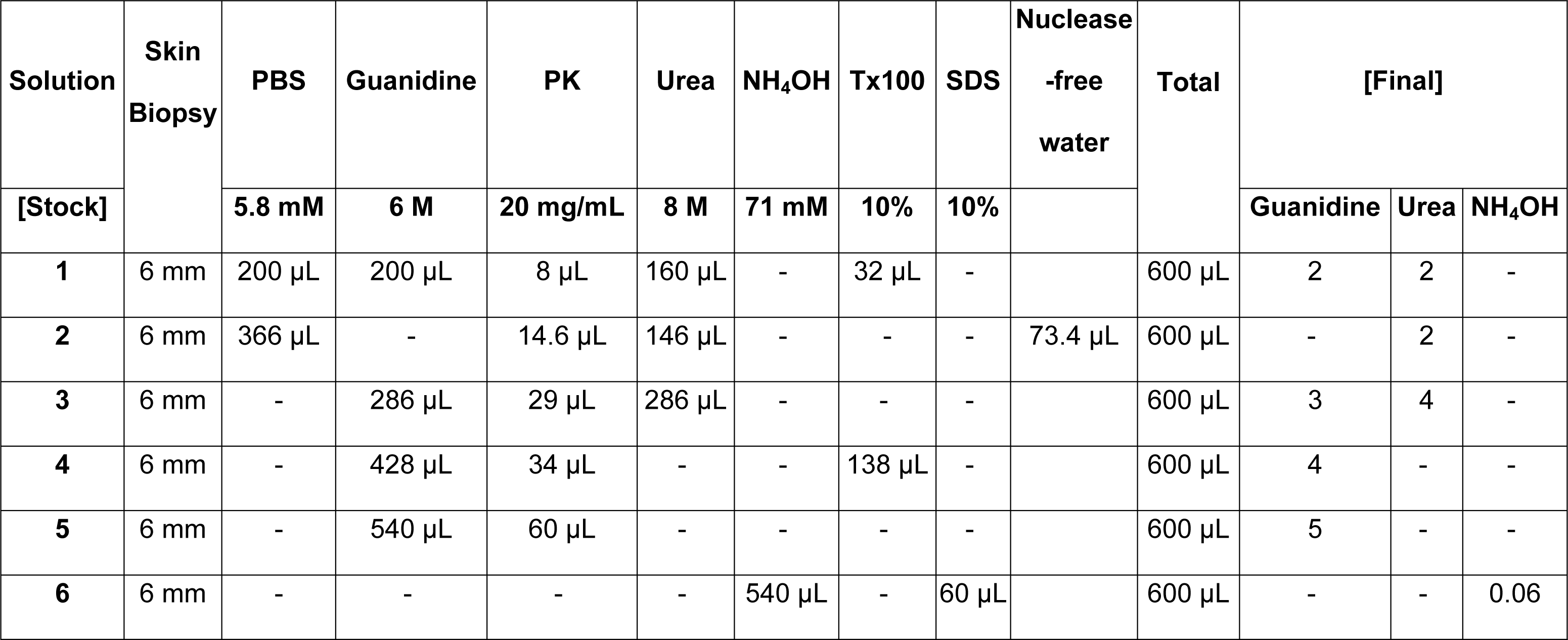
– Lysis solutions were assessed in protocols aimed at developing a simplified extraction method. Shown are the protocols and reagents with their respective concentrations and volumes necessary for the preparation of lysis solutions evaluated in DNA extraction from 6 mm skin biopsies.

Fragments of porcine skin, approximately 3 mm, were placed in microtubes containing each of the evaluated lysis solutions (TABLE 2). Subsequently, each sample was gently macerated using a sterile microtubes pestle and incubated at 56 °C for 30 minutes in a heating block, with additional maceration and vortex agitation every ten minutes. Subsequently, 200 µL of the supernatant was deposited onto FTA Elute Micro Card TM (Flinders Technology Associates – FTA, GE Whatman, Maidstone, Kent, United Kingdom) cards and stored at room temperature (21 – 23 °C) until completely dry to proceed with the purification and elution step.

### Clinical sample (biopsy) preparation and application onto FTA Micro Elute® card

Upon establishing the optimal skin biopsy DNA extraction protocol, a subsequent phase of clinical sample assessment was undertaken. To refine and appraise the streamlined protocol, the biopsy specimen was introduced into a 1.5 mL microtube, containing 146 µL of denaturing solution (8 M urea solution), 14.6 µL of proteinase K (20 mg/µL) (Roche, Germany), 366 µL of phosphate-buffered saline (PBS) (5.8 mM), and 73.4 µL of nuclease-free water (SOLUTION 2–TABLE 2). Following this, employing a sterile microtube pestle, the sample was meticulously macerated and then subjected to a 30-minute incubation at 56 °C within a thermal block. Additional rounds of maceration and vortex agitation were performed at ten-minute intervals. Subsequently, 200 µL of the resulting supernatant was meticulously deposited onto an FTA Elute Micro Card^TM^, where it was diligently preserved at ambient temperature (21 – 23 °C) until complete desiccation.

### DNA elution protocols

Ten elution protocols were evaluated using clinical skin biopsy samples with lysis solution (urea solution) stored on FTA cards (FIGURE 1). The first parameter assessed was the size of the FTA fragment to be punched out: both 3 mm and 6 mm fragments were evaluated. Subsequently, direct elution by TE buffer (pH 8.0) and the implementation of a washing step were examined. The commercial FTA wash buffer (200 µL, Qiagen, Germany) and nuclease-free water (500 µL) were tested for washing. Finally, the addition of 50 µL and 100 µL of elution buffer TE (pH 8.0), followed by incubation at 95 °C for 5 min, 10 min and 15 min were evaluated. The eluate containing the extracted DNA obtained with each protocol was subjected to a qPCR reaction to detect the *16S rRNA* and RLEP targets of *M. leprae* and the human *18S rRNA* internal control using the NAT Hans kit (IBMP, Brazil).

**Fig 1.**
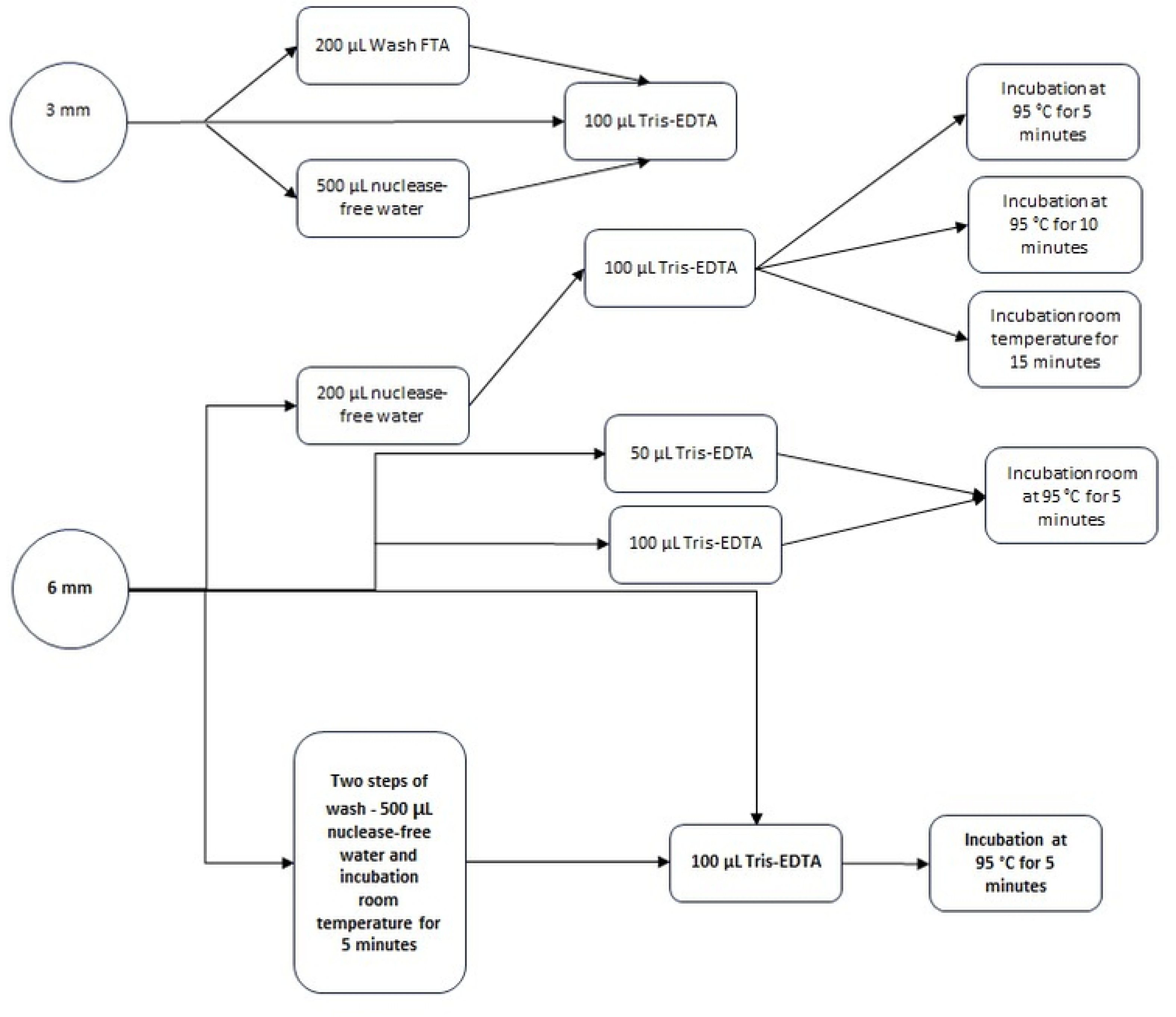
Flowchart displaying the elution protocols from FTA cards evaluated in the extraction of *Mycobacterium leprae* DNA from skin biopsy samples

### Experimental conditions for qPCR multiplex in a portable instrument (Q3-Plus)

The reactions for detection of *M. leprae* targets (*16S rRNA* and RLEP) and the human internal control (*18S rRNA*) were performed on the Q3-Plus instrument used the GoTaq Probe qPCR Master Mix (2X) (Promega, USA) and the same oligonucleotides described by Manta et al. (2022), which are used in the NAT Hans kit. For the *16S rRNA* target at concentrations of 0.75 µM for the forward and reverse oligonucleotides, and 0.3 µM for the probe; for the RLEP target, 0.4 µM for the forward and reverse oligonucleotides, and 0.2 µM for the probe; and the *18S rRNA* target, 0.2 µM for both forward and reverse, and 0.1 µM for the probe. Despite this, due to the specificities of the portable instrument, it was necessary to replace the Cy5 fluorophore with ROX, which is the probe used for the detection of the *18S rRNA* gene. On the portable platform, the reactions were standardized to a final volume of 5 µM, containing 2 µM of DNA.

Optical parameters for the FAM channel were exposure time 1 second, gain 14, and light power 8; for the VIC channel, parameters were exposure time 1 second, gain 14, and light power 9; finally, for the ROX channel, optimized parameters were exposure time 1 second, gain 15, and light power 7. The baseline was defined automatically by the instrument software (Q3-Plus V2 Suite, version 4.0, ST Microelectronics), and the established threshold was 36 arbitrary units (a.u.) for the target *16S rRNA*, 150 u.a. for RLEP and 21 u.a. for *18S rRNA* defined according to fluorescence amplitude patterns observed at different concentrations of gBlock, from *M. leprae* genome-equivalents, negative controls, and pre-characterized clinical samples.

Cycling conditions were 95 °C for 2 minutes, 45 cycles of 95 °C for 15 seconds, and 64 °C for 1 minute. The total reaction time on the portable equipment was approximately 1 hour and 40 minutes.

### Statistical analysis

Diagnostic parameters such as reaction’s efficiency, sensitivity, specificity, reproducibility, and repeatability were evaluated for the best experimental protocol using qPCR carried out on both instruments (QuantStudio-5 and Q3-Plus). The detection limit with 95% confidence interval (LOD_95%_) was calculated using the Probit model using RStudio version 4.1.0 software (downloaded from http://www.Rproject.org/). The evaluation of the agreement between the different instruments was carried out using the Bland-Altman method (ALTMAN & BLAND. 1983).

This study follows the STARD guidelines for reporting diagnostic accuracy studies (COHEN et al., 2016). The minimum information for publication of quantitative real-time PCR experiments (MiQE) (BUSTIN et al., 2009), as well as the STARD checklist, are presented as Supplemental Tables S2 and S3.

## Results

### Optimization of reactions for detection of *M. leprae* DNA in the portable instrument

#### Optical parameters and efficiency of reactions

Different values of optical parameters were evaluated in the optimization of reactions in the Q3-Plus instrument (TABLE 1). The alterations sought to increase the sensitivity and efficiency in the reactions by the instrument, through the greater amplitude of fluorescence. For FAM exposure time 1, gain 14, and light power 8 was set, while for the channel VIC the parameters were exposure time 1, gain 14, and light power 9, and for the channel ROX the optimized parameters were exposure time 1, gain 15, and light intensity 7.

The fluorescence threshold for each target was set at 36 a.u. for the *16S rRNA* target (FAM), 150 a.u. for the RLEP target (VIC) of *M. leprae*, and 21 a.u. for the *18S rRNA* target (ROX), the human internal control.

The efficiency was calculated based on a dilution curve (10^5^ – 10^0^ copies/μL) of synthetic DNA (gblock) results. For Q3-Plus equipment the efficiency was 109% for the *16S rRNA* gene and 108% for the RLEP target. The standard QS-5 equipment showed 102% efficiency for *16S rRNA* and 94% for RLEP (FIGURE 2).

**Fig 2.**
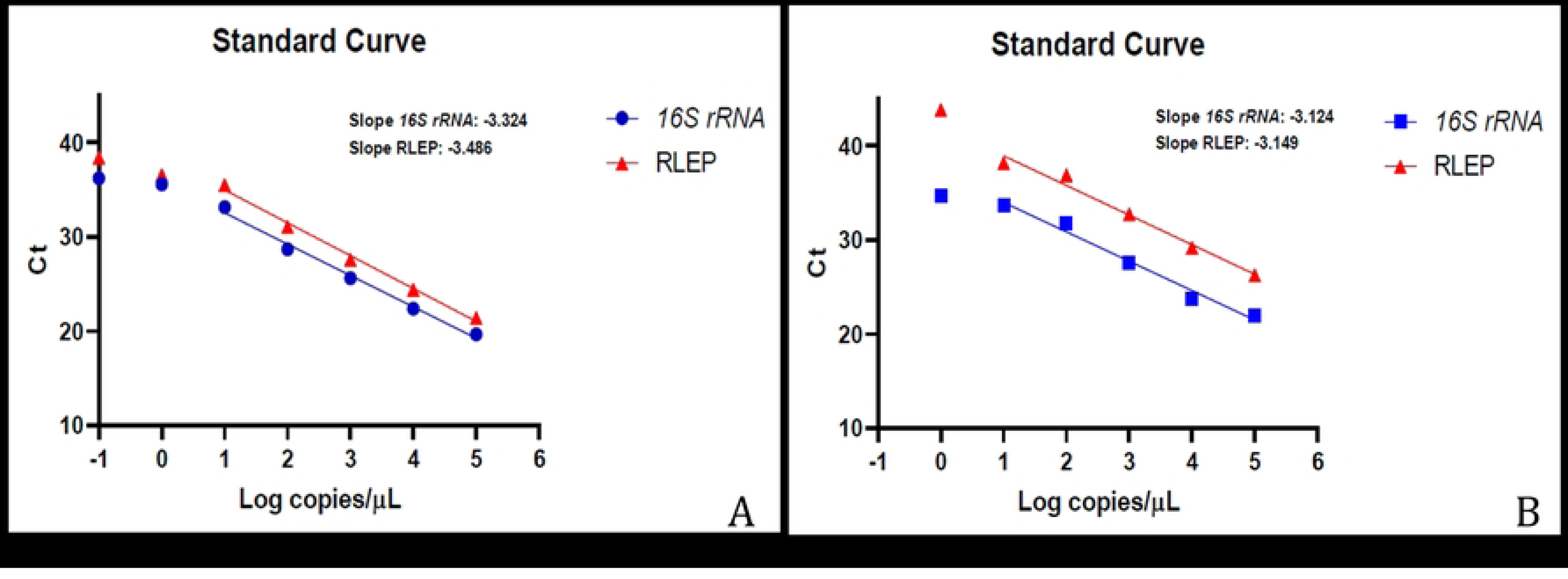
Standard curve of qPCR reactions analyzed by QuantStudio and Q3-Plus equipment for the detection of *Mycobacterium leprae 16S rRNA* and RLEP targets from synthetic DNA (A) Reactions on QuantStudio 5; (B) Reactions on portable platform Q3-Plus. Linear regressions were obtained from no less than 4 independent experiments

The same analyzes were performed from reactions with DNA samples of the genome-equivalent to 10^6^ cells/µL of *M. leprae*. The efficiency of 131% for the reactions of the *16S rRNA* target and 105% for the RLEP target were observed in the Q3-Plus equipment, and an efficiency was 101% for *16S rRNA* and 100% for RLEP in the QS-5 equipment (FIGURE 3).

**Fig 3.**
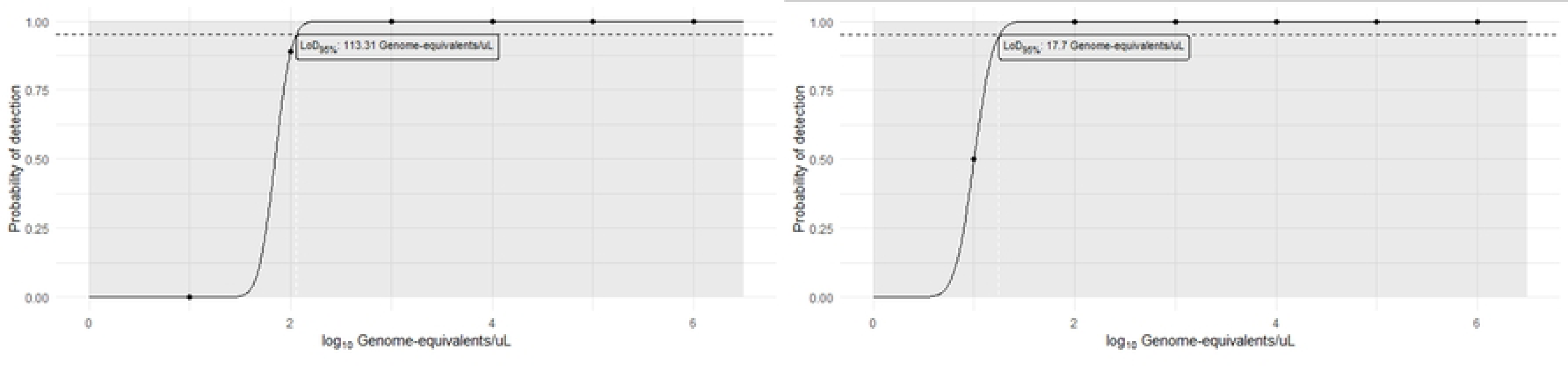
Standard curve of qPCR reactions analyzed by QuantStudio and Q3-Plus equipment for the detection of *Mycobacterium leprae 16S rRNA* and RLEP targets using purified DNA from *M. leprae* (A) Reactions on QuantStudio 5; (B) Reactions on portable platform Q3-Plus. Linear regressions were obtained from no less than 4 independent experiments.

#### Analytical sensitivity

The limit of detection (LOD_95%_) on the Q3-Plus equipment using synthetic DNA was 13.86 copies/μL for the *16S rRNA* and RLEP targets. On the QS-5 equipment, the respective values were found to be 12.45 copies/μL for *16S rRNA* and 20.44 copies/μL for RLEP. When using purified *M. leprae* DNA, the LOD_95%_ on the Q3-Plus instrument was 113.31 genome-equivalents/μL for the *16S rRNA* gene and 17.70 genome-equivalents/μL for the RLEP (Fig 4). On the standard equipment (QS-5), the values were 205.26 genome-equivalents/μL for *16S rRNA*, and 15.34 genome-equivalents/μL for the RLEP.

**Fig 4.**
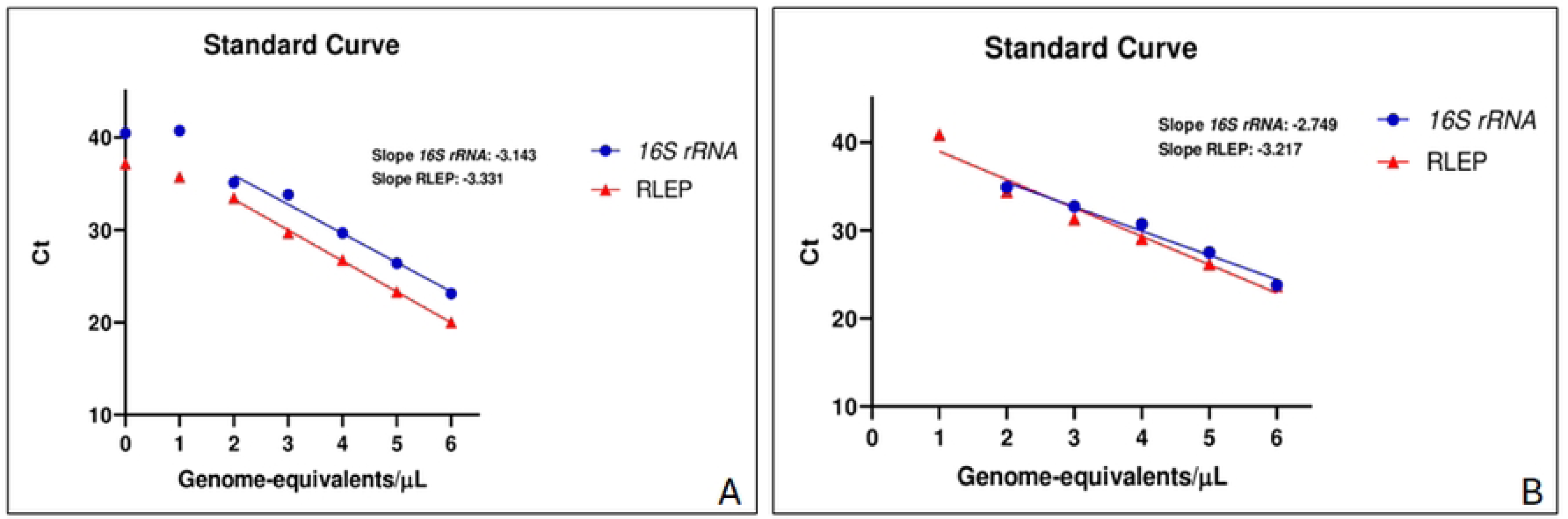
Analytical sensitivity of qPCR reactions for the targets *16S rRNA* and RLEP of Mycobacterium leprae performed on the portable Q3-Plus equipment. (A) LOD95% 16S rRNA; (B) LOD95% RLEP.

#### Reproducibility and repeatability in the portable equipment

Results obtained in the intra– and inter-operator replicate series are shown in TABLE 2 and show the coefficients of variation observed in the intra-operator reactions at the lowest concentrations (10^1^ – 10^0^ copies/μL). Although these values are greater than 5%, they are lower than 10%, thus being non-significant for both the *16S rRNA* and RLEP targets. In the inter-operator analyses the coefficient of variation values between concentrations was between 0.05 and 3.09% for the *16S rRNA* gene. For the RLEP target, values ranged from 0.55 to 3.10%. The analysis of variance (ANOVA) performed for the results obtained by the different operators, assuming a confidence interval of 95%, corroborated the previous analyses, showing no significant inter-operator difference (p-value = 0.994 for *16S rRNA*, and p-value = 0.992 for RLEP) (TABLE 3).

**TABLE 3.** – Repeatability and reproducibility analyses, including the respective coefficients of variation, were determined in qPCR reactions targeting *Mycobacterium leprae 16S rRNA* and RLEP on the Q3-Plus equipment.

| Target | Synthetic DNA | Repeatability |  |  |  |  |  |  |  |  |  |  |  | Reproducibility |  |  |  |
| --- | --- | --- | --- | --- | --- | --- | --- | --- | --- | --- | --- | --- | --- | --- | --- | --- | --- |
|  |  | Operator 1 |  |  |  | Operator 2 |  |  |  | Operator 3 |  |  |  | Interoperator |  |  |  |
|  | Copies/µL | mean | desv | desv pad | %rRSD | mean | desv | desv pad | %rRSD | mean | desv | desv pad | %rRSD | mean | desv | desv pad | %rRSD |
| Mean Ct<br>16S<br>rRNA | 1,00E+04 | 22.83 | 0.25 | 0.32 | 1.41 | 22.78 | 0.26 | 0.35 | 1.55 | 22.66 | 0.10 | 0.15 | 0.68 | 22.76 | 0.06 | 0.09 | 0.39 |
|  | 1,00E+03 | 26.61 | 0.11 | 0.15 | 0.55 | 26.60 | 0.12 | 0.16 | 0.61 | 26.62 | 0.07 | 0.09 | 0.35 | 26.61 | 0.01 | 0.01 | 0.05 |
|  | 1,00E+02 | 30.05 | 0.30 | 0.39 | 1.31 | 29.75 | 0.14 | 0.19 | 0.65 | 30.16 | 0.30 | 0.43 | 1.43 | 29.99 | 0.16 | 0.21 | 0.71 |
|  | 1,00E+01 | 33.71 | 0.91 | 1.18 | 3.49 | 34.07 | 1.34 | 1.97 | **5.80** | 34.40 | 0.60 | 0.60 | 1.73 | 34.06 | 0.23 | 0.34 | 1.01 |
|  | 1,00E+00 | 34.65 | 0.51 | 0.74 | 2.13 | 36.47 | 1.14 | 1.44 | 3.96 | 36.67 | 2.39 | 3.39 | **9.23** | 35.93 | 0.85 | 1.11 | 3.09 |
| Mean Ct<br>RLEP | 1,00E+04 | 28.77 | 0.23 | 0.32 | 1.13 | 29.19 | 0.96 | 1.27 | 4.34 | 28.56 | 0.51 | 0.68 | 2.38 | 28.84 | 0.23 | 0.32 | 1.11 |
|  | 1,00E+03 | 31.84 | 0.21 | 0.28 | 0.87 | 32.36 | 0.61 | 0.80 | 2.46 | 32.03 | 0.18 | 0.24 | 0.76 | 32.07 | 0.19 | 0.26 | 0.82 |
|  | 1,00E+02 | 35.20 | 0.26 | 0.34 | 0.97 | 34.90 | 0.25 | 0.35 | 1.00 | 35.30 | 0.51 | 0.67 | 1.91 | 35.13 | 0.15 | 0.21 | 0.59 |
|  | 1,00E+01 | 38.75 | 0.92 | 1.07 | 2.75 | 38.72 | 1.24 | 1.61 | 4.17 | 39.10 | 0.17 | 0.26 | 0.66 | 38.86 | 0.16 | 0.21 | 0.55 |
|  | 1,00E+00 | 40.44 | 1.46 | 2.06 | **5.10** | 38.11 | * | * |  | 38.70 | 0.94 | 1.33 | 3.44 | 39.08 | 0.91 | 1.21 | 3.10 |

#### Evaluation of the portable qPCR using DNA from skin biopsies

A difference in the Ct value was observed for the same 95 samples extracted by commercial kit when analyzed on different instruments. The mean Ct difference for the same samples between the instruments resulted in an increase of 1.40 cycles for the Q3-Plus instrument to *16S rRNA* target and an increase of 5.15 cycles for the RLEP target when compared to that obtained in the standard instrument analysis (Supplemental Figure S1 and S2). In the internal control (*18S rRNA*) there was a mean increase of 1.42 cycles on Q3-Plus analysis (Supplemental Figure S3). Cycle **v**alues above these means were mainly observed in samples classified as PB, where a lower concentration of the target DNA is expected.

To determine the cutoff to reactions on Q3-Plus, the mean difference observed between the instruments for the different targets was added to the values established for the NAT Hans kit. In this protocol, the cutoff values for targets are Ct 35.5 for *16S rRNA;* and Ct 34.5 for RLEP. Therefore, for the reactions analyzed in Q3-Plus the Ct cutoffs established for the *16S rRNA* target was 36.9, and for the RLEP target it was 39.6. Consequently, for samples that exhibited amplification for both targets, with values below 36.9 and 39.6 for the *16S rRNA* and RLEP, respectively, they were classified as positive on the Q3-Plus equipment. For those samples that displayed Ct values above the cutoff point or absence of amplification, a classification of negative assigned. Moreover, samples that exhibited amplification in only one of the targets (*16S rRNA* or RLEP) were classified as indeterminate. In these cases, a retest of the sample and patient follow-up is recommended. In this study, of all 95 samples analyzed, 43% (41/95) were considered qPCR positive for the presence *M. leprae* DNA, 43% (41/95) negative, and 14% (13/95) indeterminate.

On the standard equipment (QS-5), the same samples evaluated using the NAT Hans kit were classified as positive, negative, or indeterminate according to the recommendation cutoff of the commercial kit protocol. Following the analysis, 47% (45/95) of the samples were determined as positive, while 40% (38/95) were negative for the agent. The remaining 13% (12/95) of the samples were classified as indeterminate.

The NAT Hans reactions on Q3-Plus equipment demonstrated a sensitivity of 73% and specificity of 85%. However, out of the 95 samples analyzed, 14% (13/95) yielded indeterminate results. The same reactions conducted on the standard equipment (QS-5) showed a sensitivity of 77%, specificity of 77%, and 13% (12/95) of samples with indeterminate results (TABLE 4). The positive predictive values for the Q3-Plus equipment were 88%, and for the QS-5, it was 82%. The negative predictive values were 68% and 71% for Q3-Plus and QS-5 equipment, respectively. Regarding the accuracy of the tests, it was 78% for Q3 equipment and 77% QS-5.

**TABLE 4.** – Comparison of molecular diagnostic parameters in clinical biopsies extracted by commercial protocol and evaluated in ABI7500, QuantStudio-5 and Q3-Plus platforms.

| Parameters | Commercial extraction |  |
| --- | --- | --- |
|  | Quantstudio 5 | Q3-Plus |
| Sensitivity | 77% | 73% |
| Specificity | 77% | 85% |
| Accuracy | 77% | 78% |
| PPV | 82% | 88% |
| NPV | 71% | 68% |

## Analysis of extractions protocols

### Evaluation of Lysis Solutions for Porcine Skin Models

Several different lysis solutions devised to simplify the DNA extraction process from skin samples were evaluated visually and by qPCR using porcine skin as model tissue (HWANG et al., 2021; SUMMERFIELD et al., 2015). TABLE 5 summarizes all six protocols that were evaluated and the corresponding mean Ct for qPCR detection of the mammalian *18S rRNA* gene. A mixture consisting of urea (2 M), proteinase K (0.5 mg/mL), and PBS pH 7.4 (3.5 mM) yielded the best results in the visual evaluations regarding the turbidity of the solution and reduction/dissolution of the fragment of skin (FIGURE 5), and in the detection of the *18S rRNA* gene by qPCR in terms of fluorescence amplitude and Ct. In the porcine tissue used as a comparative model the mean Ct was 18.74 (range 17.45 to 19.40) in the detection of the *18S rRNA* gene.

**FIGURE 5.**
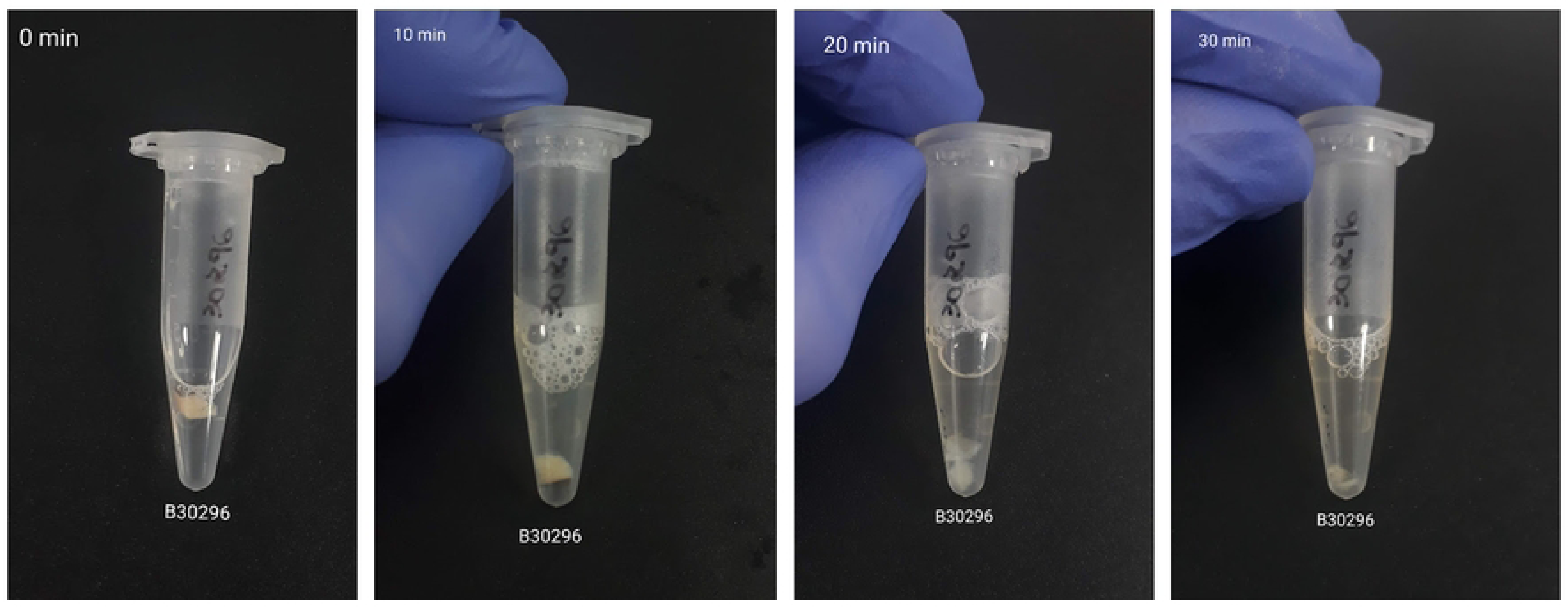
Demonstration of the simplified extraction protocol in clinical biopsy sample. After each 10-minute step, a reduction in skin fragments and a change in the turbidity of the solution were observed.

**TABLE 5.** – Results of Ct and average Cts obtained from evaluations for definition of the lysis solution for each simplified extraction protocol, derived from the detection of the mammalian *18S rRNA* target by qPCR in porcine skin samples. Protocol numbers are summarized as 1 – Guanidine (2 M) and Urea (2 M); 2 – Urea (2 M); 3 – Guanidine (3 M) and Urea (4 M); 4 – Guanidine (4 M); 5 – Guanidine (5 M); 6 – NH_4_OH (0.06 M); NC – Negative control.

| Protocols | 1 | 2 | 3 | 4 | 5 | 6 | NC |
| --- | --- | --- | --- | --- | --- | --- | --- |
|  | 30.75 | 21.87 | 23.69 | 24.77 | 28.11 | 23.93 | 29.50 |
| **Ct** | 26.57 | 21.16 | 23.11 | 23.03 | 28.81 | 31.68 | 29.78 |
|  | 29.08 | 22.08 | 24.26 | 23.62 | 27.76 | 25.01 | 29.27 |
| **Mean Ct** | **28.80** | **21.70** | **23.69** | **23.81** | **28.23** | **26.87** | **29.52** |

### Evolution of elution protocols

Among the various elution protocols, the best outcomes, also assessed through amplification of the *18S rRNA* gene, were achieved using the 6 mm puncher, along with two washing steps employing 500 µL of nuclease-free water, and subsequent incubation with 100 µL of TE (pH 8.0) at 95 °C for 5 minutes in a thermal block. A schematic representation of the final comprehensive protocol is illustrated in FIGURE 6.

**FIGURE 6.**
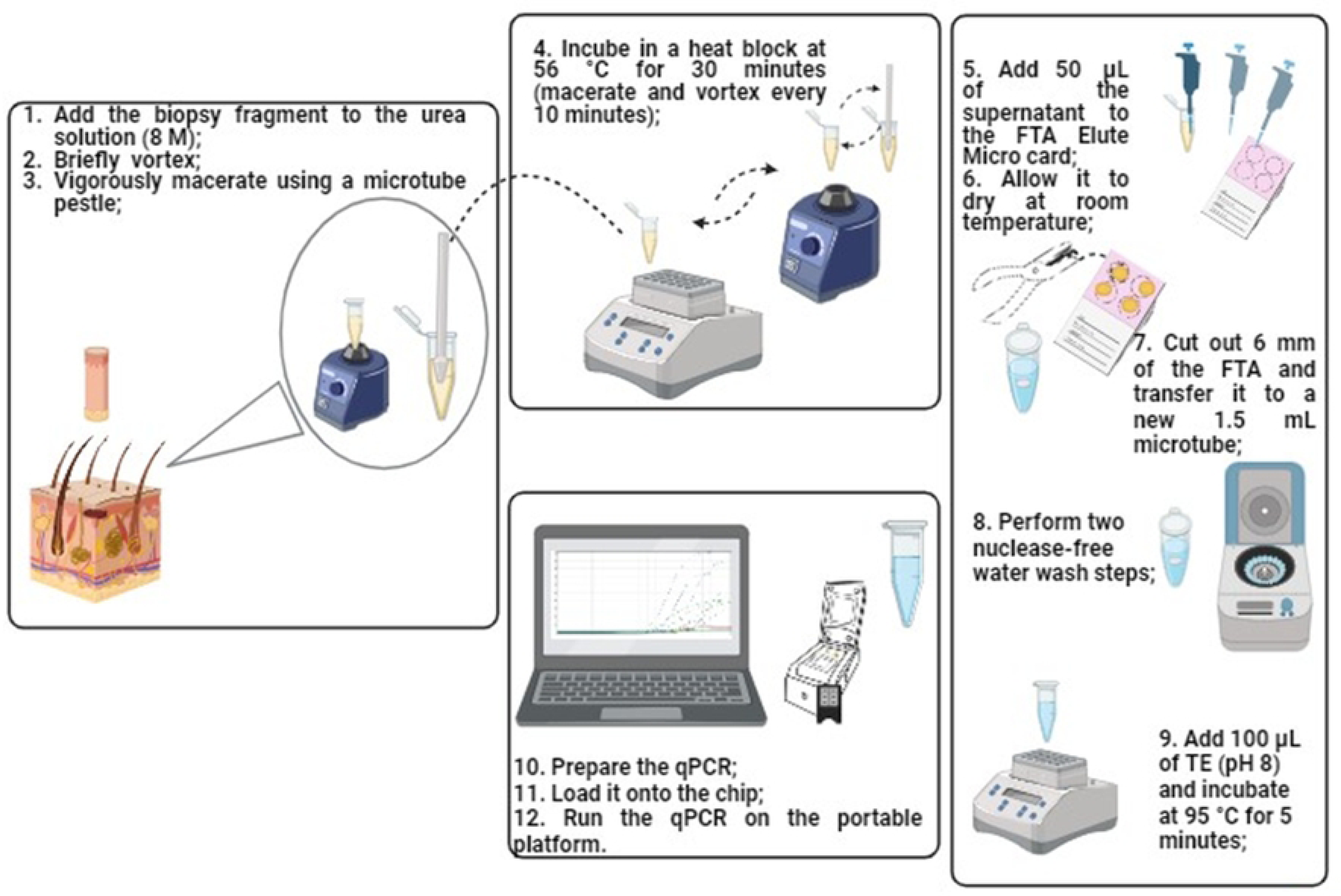
Schematic representation of the simplified DNA extraction protocol for *Mycobacterium leprae* from clinical skin biopsy samples. (Credit: Created in BioRender.com).

### Analysis of clinical samples (skin biopsy) using a developed extraction protocol and portable platform (Q3-Plus)

The entire methodology developed, from DNA extraction using the simplified extraction protocol (SOLUTION 2 – TABLE 2) to qPCR analysis for the detection of *M. leprae* targets on the portable platform, was evaluated using clinical skin biopsy samples from patients with leprosy (MB and PB) and patients with other dermatoses (OD).

During visual assessment while extracting genetic material, a reduction in the skin biopsy fragment or its complete dissolution was observed, resulting in a visibly altered solution turbidity. Successful detection of the *18S rRNA* gene was also achieved. In reactions conducted on the standard equipment (QS-5), the mean Ct was 21.12 (ranging from 17.24 to 28.43), while on the Q3-Plus equipment, the mean Ct was 22.46 (ranging from 19.02 to 30.54).

In a Bland-Altman test conducted based on the same group of samples, with 95% confidence the *16S rRNA* target, presented the mean threshold cycle variation between paired samples was 1.29 cycles above on the Q3-Plus equipment (Supplemental Figure S4). For the detection of the *M. leprae* RLEP target, a mean variation of 4.44 threshold cycles above was observed for the Q3-Plus equipment (Supplemental Figure S5). Finally, for the 18S rRNA, a mean variation of 1.34 threshold cycles was observed (Supplemental Figure S6).

The qPCR reaction using the NAT Hans kit on the Q3-Plus equipment yielded positivity for *M. leprae* in 26% (14/53) of the samples, while 57% (30/53) of the samples tested negative for the agent, and 17% (9/53) of samples with indeterminate results.

For those analyses conducted on the standard equipment (QS-5), 30% (16/53) tested positive for the *M. leprae* agent, while 66% (35/53) yielded negative results. On this equipment, 4% (2/53) of the samples produced indeterminate results.

As for the sensitivity and specificity parameters of the testes, the portable platform (Q3-Plus) exhibited 52% and 87%, respectively. The reaction analyzed by the standard equipment (QS-5) showed 64% sensitivity and 93% specificity. Regarding the accuracy of the tests, it was 70 % for the Q3-Plus equipment and 80% for the QS-5 equipment. The positive predictive value (PPV) for the analyses conducted on the Q3-Plus equipment was 79%, whereas on the QS-5 equipment it was 88%. The negative predictive value (NPV) was 67% and 77% for the Q3-Plus and QS-5 equipment, respectively (TABLE 6).

**TABLE 6.** – Comparison of molecular diagnostic parameters in clinical biopsies extracted using both commercial and simplified protocols across different platforms.

| Parameters | Commercial extraction<br>ABI7500 | Simplified protocol<br>Quantstudio-5 | Simplified protocol<br>Q3-Plus |
| --- | --- | --- | --- |
| Sensitivity | 74% | 64% | 52% |
| Specificity | 65% | 93% | 87% |
| Accuracy | 69% | 80% | 70% |
| PPV | 70% | 88% | 79% |
| NPV | 69% | 77% | 67% |

The standard analysis (commercial extraction method and standard thermocycler instrument) in the same group of the samples presented 38% (20/53) positives, 30% (16/53) negatives, and 32% (17/53) indeterminate results. Parameters of sensibility and specificity demonstrate 74% and 65%, respectively. Accuracy was estimated at 69%. Positive predictive value and negative predictive value were 70% and 69%, respectively.

## Discussion

The utilization of screening tests in settings with restricted resources, along with their contribution to active case detection, holds paramount significance in achieving reduced leprosy incidence rates (BRASIL, 2022; STEINMANN et al., 2017). Considering the absence of a definitive gold standard test, the inherent constraints of adjunctive assays, and the clinical nuances of the disease, the advancement of assays characterized by high sensitivity and specificity on portable platforms, accompanied by cost reduction and technique streamlining, facilitates the adoption of preventive and control interventions within the disease transmission continuum.

In the present study, the optimization of qPCR reactions on a portable analysis platform (Q3-Plus) was established using the oligonucleotides included in the first national diagnostic kit for leprosy, approved by ANVISA (kit NAT Hans – IBMP). Being a point-of-care device due to its compact dimensions, reaction volumes were optimized to 5 µL, requiring minor adjustments to oligonucleotide concentrations and master mix composition. Nevertheless, the Q3-Plus exhibited excellent efficiency values, closely approaching those of the standard equipment Quantstudio-5, highlighting the applicability of the platform in analytical contexts.

The efficiency of reaction is linked to the exponential amplification of the target material throughout the analysis. Factors such as the purity and concentration of the target in the sample and reagents, as well as the final volume of the qPCR reaction, are known determinants of the technique’s efficiency (SVEC et al., 2015). Although the final reaction volume in the Q3-Plus equipment is five times smaller than that used in reactions analyzed in the standard equipment, the efficiency values remained close to 100%, which is desirable for this type of analysis (SVEC et al., 2015). Previous studies on reaction optimization for molecular detection of *Plasmodium* spp., *Trypanosoma cruzi*, and *Mycobacterium tuberculosis* had already noted higher efficiencies in the portable device when compared to the standard (ABI7500) (ALI et al., 2020; RAMPAZZO et al., 2019), as observed in the current study. This reinforces the possibility of this parameter being an anticipated trait of the Q3-Plus equipment.

The determination of the fluorescence threshold is an important tool to ensure that non-specific amplifications or other interferents do not generate false results in molecular qPCR tests. This parameter can be determined through numerical analysis or visually by the operator (CARAGUEL et al., 2011). Despite its subjectivity, in the present study, this parameter was manually defined by the operator through the observation of the fluorescence amplitude in known samples.

The reactions from synthetic DNA (gBlock^®^) in both instruments exhibited linearity up to 10^1^ copies/µL, and beyond this range, amplifications started to occur stochastically. In the analyses of equivalent genome/µL of *M. leprae*, linearity in the instruments extended up to 10^2^ equivalent genomes/µL; however, beyond this range, the Q3-Plus instrument lost analytical sensitivity for *16S rRNA* target, while the QS-5 began to exhibit random amplifications. For the RLEP target, amplifications persisted beyond the linear range in both instruments. Given that this is a multicopy target, higher analytical sensitivity is expected (COLE et al., 2001). The loss of reaction linearity implies the occurrence of random amplifications, which could interfere particularly in cases classified as paucibacillary due to low bacillary load (MARTINEZ et al., 2014).

Through the analysis of the limit of detection (LOD_95%_), it was possible to confirm greater sensitivity for the RLEP target. Reactions with purified *M. leprae* DNA yielded LOD values of 113.31 genome-equivalents/µL for the *16S rRNA* gene and 17.70 genome-equivalents/µL for RLEP on the Q3-Plus instrument. In a previous study by MANTA et al. (2020), the authors reported LOD values of 126 genome-equivalents/reaction for the *16S rRNA* and 1.3 genome-equivalents/reaction for the RLEP target, using the NAT Hans kit in a standard thermocycler (ABI7500). This proximity of values ensures the good analytical sensitivity of the portable instrument. It is important to emphasize that the difference in LOD_95%_ values between the targets is expected, as RLEP is a repetitive element in the genome with approximately 36 copies (MANTA et al., 2022; COLE et al., 2001).

Analysis intra and inter-operator show very good results. All coefficients of variation were found to be below 5%. The three data points from intra-operator assessments that exceed this variation might be explained as they correspond to the lowest concentrations of the target DNA, falling outside or at the limit of the reaction, where the probability of amplification decreases and becomes stochastic.

In Bland-Altman analyses on pre-characterized clinical samples, a mean variation of approximately 1.40 cycles higher for the *16S rRNA* target was observed on the Q3-Plus equipment compared to QS-5. For the RLEP target, the observed variation was approximately 5.15 cycles higher in the Q3-Plus evaluated samples. These values closely align with those reported by RAMPAZZO et al. (2019) from the optimization of the Q3 equipment for molecular detection of *T. cruzi* and *Plasmodium* spp. The disparity noted by these authors amounted to an increase of 2 to 4 Cts in Q3-Plus reactions.

The results obtained from optimization of reactions on the portable platform, concerning optical parameters, reaction efficiency, and analytical sensitivity, were confirmed in pre-characterized clinical samples extracted using the commercial kit (Qiagen). Analyses of sensitivity, specificity, and accuracy comparable to those of established qPCR tests on the standard equipment confirm the applicability of point-of-care testing. The occurrence of false negatives is observed especially in paucibacillary cases (MARTINEZ et al., 2011; ROSA et al., 2013). The low bacterial load hinders the detection of these cases. However, qPCR is still considered the best technique to be used as a screening test complementary diagnostic due to its high sensitivity and specificity, particularly in detecting PB cases (MARTINEZ et al., 2011; WICHITWECHKARN et al., 1995).

Among the evaluated extraction protocols, certain chaotropic agents such as urea, guanidine, and ammonium hydroxide (NH_4_OH) were considered. In this study the FTA cards aided in the isolation and purification steps of the genetic material. Due to their affinity for the cellulose fibers in the card, DNA recovery was feasible after the washing steps (DAIRAWAN & SHETTY, 2020).

FTA cards (Whatman^®^) are composed of cellulose fibers or other materials with affinity for genetic material (DNA). Their composition may include reagents capable of assisting in the cellular lysis and protein denaturation steps, such as sodium dodecyl sulfate (SDS) and sodium lauroyl sarcosinate (SLS), facilitating DNA exposure. They were developed to streamline sample transport and storage at room temperature, while ensuring DNA viability for molecular analyses of interest (BURGOYNE et al., 2003; AYE et al., 2011). They require minimal space for storage and have low risk of cross-contamination (SANTOS, 2018).

Regarding the application of urea solution (8 M) for DNA extraction from tissue samples, the results concerning the enhanced dissolution of skin fragments may be linked to the improved activity of proteinase K facilitated by high urea concentrations. Additionally, urea may have contributed to the preservation of the obtained genetic material, as reported in previous studies (AHMED, 1993; HILZ et al., 1975). Improved dissolution quality was also observed through qPCR for the human *18S rRNA* target, as the Ct values were earlier than those found in extractions using other protocols and more consistent across replicates.

Regarding the analyses for *M. leprae* targets (*16S rRNA* and RLEP), there was a loss of sensitivity in the reactions from the simplified protocol-extracted samples, as evidenced by the increased Ct value in the *16S rRNA* target when compared to the standard test performed by kit NAT Hans in samples extracted by commercial method (Qiagen).. Therefore, it is suggested that greater interference may be linked to the quality of genetic material extraction from the bacillus or residual extraction components that were not adequately removed during the purification/washing steps.

This decrease in sensitivity affects the detection of paucibacillary cases, as observed in the study samples. The reported sensitivity range varies (36.4% to 85%) in studies using qPCR for different targets and biological materials. In these studies, lower sensitivity is also observed for PB cases, demonstrating the intrinsic limitation in detecting this clinical form (BRAET et al.,2021; BARBIERI et al., 2019; MANTA et al., 2019; CHENG et al., 2019; AZEVEDO et al., 2017).

The results of the present study demonstrate the need for further protocol optimization to improve the detection of PB cases. However, the results presented are promising. The complete technological platform can serve as an auxiliary tool in detecting leprosy cases in remote regions and vulnerable populations. Social vulnerability, particularly observed in areas with low infrastructure, is relevant in perpetuating the disease transmission chain (de SOUZA et al., 2019; CABRAL-MIRANDA et al., 2014).

Furthermore, the possibility of using molecular tests may reduce recurring misdiagnoses in leprosy (NEVES et al., 2023; MANTA et al., 2020). Ensuring diagnosis for all populations is essential, and decentralizing access to it, as facilitated by active case finding, is pivotal for leprosy to discontinue being considered a public health problem in Brazil (BARBIERI et al., 2016). The Global Leprosy Strategy 2021-2030, published by the World Health Organization (WHO) (WHO, 2021), aims to eliminate the disease by interrupting transmission. However, there is a consensus that this goal will only be achieved with the improvement of the current strategies of complementary diagnosis, with an emphasis not only on developing strategies to increase the sensitivity of current tests, but also to increase access to available tests. The portable platform serves as a tool that, by adhering to the principles of the point-of-care testing concept, can contribute to overcoming some of the limitations in leprosy diagnosis.

As a limitation of this study, it should be considered that the samples were collected based on their occurrence in the clinic. The final clinical outcome of cases will be determined after one year of follow-up. Additionally, molecular test positivity can also occur in cases under treatment, where residual bacillus DNA may be present.

## Conclusion

The optimization of qPCR reactions on the portable Q3-Plus platform for aiding leprosy diagnosis has shown promising potential for the full application of this technology as an auxiliary tool for healthcare professionals in suspected cases. Concerning the evaluated lysis solutions, the urea solution (2 M) demonstrated the best outcomes both visually, regarding fragment dissolution and alteration of medium turbidity, and through qPCR assessment for the detection of the human *18S rRNA* gene.

The complete technological solution (DNA extraction using a simplified protocol and qPCR analysis on the portable platform) yielded promising results. However, concerning the test’s sensitivity, further optimization will be needed to enhance the detection of paucibacillary cases.

## Data Availability

All data underlying the findings are contained within the manuscript.

## Acknowledgments

The authors are grateful to the entire team of dermatologists, nurses and technicians that collaborate at the Souza Araújo Clinic from the Leprosy Laboratory at the Oswaldo Cruz Institute. We especially thank Raquel Barbieri, Alexsandro Cruz Barreto and Cristiane Domingues for all the technical and administrative assistance.

This study was supported by grants from Carlos Chagas Filho Research Support Foundation of the State of Rio de Janeiro (E-26/203.913/2022) Carlos Chagas Institute Research Stimulus Program (ICC 008 FIO 21 – SUB 22) and by the National Fund for Health/Brazilian Ministry of Health (TED 69/2021). LSD is a CNPq fellowship holder, FSNM is a Faperj fellowship holder, and ADTC is a CNPq productivity fellow (level 2).

## List of Supplementary Material

**Supplemental Table S1**. Information regarding clinical samples, gender, age and clinical diagnosis collected at the Hansen’s disease Laboratory of Oswaldo Cruz Institute – Fiocruz – RJ. Key: PB – Paucibacillary; MB – Multibacillary; OD – Other dermatoses.

**Supplemental Table S2**. MIQE checklist.

**Supplemental Table S3**. STARD checklist.

**Supplemental Figure S1**. Bland-Altman analysis for the *16S rRNA* target detected in the portable instrument versus the benchtop instrument. The mean difference it was 1.40 cycle of threshold between instruments. The upper limit of agreement with 95% confidence interval was 9.40 and the lower limit of agreement was –6.61.

**Supplemental Figure S2**. Bland-Altman RLEP (Optimization multiplex qPCR Q3-Plus). Bland-Altman analysis in RLEP target. The mean difference it was 5.15 cycle of threshold between the equipment in 95% confidence interval. The upper limit of agreement was 10.59 and the lower limit of agreement was –0.28.

**Supplemental Figure S3**. Bland-Altman *18S rRNA* (Optimization multiplex qPCR Q3-Plus). Bland-Altman analysis in *18S rRNA* target. The mean difference it was 1.42 cycle of threshold between the equipment in 95% confidence interval. The upper limit of agreement was 3.25 and the lower limit of agreement was –0.40.

**Supplemental Figure S4**. Bland-Altman *16S rRNA* (Evaluated simplified extraction protocol in clinical samples analyzed in Q3-Plus). Bland-Altman analysis in *16S rRNA* target. The mean difference it was 1.29 cycle of threshold between the equipment in 95% confidence interval. The upper limit of agreement was 12.39 and the lower limit of agreement was –9.81.

**Supplemental Figure S5**. Bland-Altman RLEP (Evaluated simplified extraction protocol in clinical samples analyzed in Q3-Plus). Bland-Altman analysis in RLEP target. The mean difference it was 4.44 cycle of threshold between the equipment in 95% confidence interval. The upper limit of agreement was 11.91 and the lower limit of agreement was –3.04.

**Supplemental Figure S6**. Bland-Altman *18S rRNA* (Evaluated simplified extraction protocol in clinical samples analyzed in Q3-Plus). Bland-Altman analysis in *18S rRNA* target. The mean difference it was 1.34 cycle of threshold between the equipment in 95% confidence interval. The upper limit of agreement was 6.90 and the lower limit of agreement was –4.22.

## Author contribution list

Specific Contributions:

1. ABS (Lead Author)
  a. Collected and analyzed the data;
  b. Drafted the initial manuscript.
2. LSD
  a. Laboratory support
3. MRA
  a. Responsible for statistical analyses
  b. Revised the manuscript;
4. RPO
  a. Revised the manuscript;
  b. Provided funding
5. FSNM
  a. Design of the study;
  b. Supervised sample collection;
  c. Performed, and analyzed same experiments;
  d. Contributed to the interpretation of results;
  e. Revised the manuscript;
6. ADTC
  a. Design of the study;
  b. Conceptualized the study
  c. Contributed to the interpretation of results;
  d. Supervised lab data collection
  e. Provided funding
  f. Critically reviewed intellectual content.
7. MOM
  a. Provided funding;
  b. Conceptualized the study

